# Pharmacological and psychological treatment have common and specific effects on brain activity in obsessive-compulsive disorder

**DOI:** 10.1101/2022.08.08.22278405

**Authors:** AL van der Straten, WB Bruin, LA van de Mortel, F ten Doesschate, MJM Merkx, PP de Koning, NCC Vulink, M Figee, OA van den Heuvel, D Denys, GA van Wingen

## Abstract

**Background:** Initial treatment for obsessive-compulsive disorder (OCD) consists of pharmacological treatment with selective serotonin reuptake inhibitors (SSRIs) and/or psychological treatment with cognitive-behavioral therapy (CBT). The assumption is that both treatments have different neural working mechanisms, but empirical evidence is lacking. We investigated whether these treatments induce similar or different functional neural changes in OCD.

**Methods:** We conducted a longitudinal non-randomised controlled trial in which thirty-four OCD patients were treated with sixteen weeks of CBT or SSRIs. Functional magnetic resonance imaging was performed before and after treatment during emotional processing (emotional face matching and symptom provocation tasks) and response inhibition (stop signal task). Twenty matched healthy controls were scanned twice with a similar time interval. The study was registered at the Netherlands Trial Registry (NTR6575), https://trialsearch.who.int/Trial2.aspx?TrialID=NTR6575.

**Results:** Both CBT and SSRIs were successful in reducing OCD symptoms. Compared to healthy controls, treatment led to a reduction of insula activity in OCD patients during symptom provocation. The comparison between treatment groups revealed wide-spread divergent brain changes in the cerebellum, posterior insula, caudate nucleus, hippocampus, occipital and prefrontal cortex during all tasks, explained by relative increases of activity following CBT compared to relative decreases of activity following SSRIs.

**Conclusions:** Pharmacological and psychological treatment primarily lead to opposite changes in brain function, with a common reduction of insula activity during symptom provocation. These findings provide insight in common and specific neural mechanisms underlying treatment response, suggesting that CBT and SSRIs support recovery from OCD along partly distinct pathways.

## Introduction

Obsessive-compulsive disorder (OCD) is a debilitating psychiatric disorder characterized by repetitive thoughts (obsessions) and repetitive behaviors or mental rituals (compulsions). Initial treatment for OCD consists of pharmacological treatment with selective serotonin reuptake inhibitors (SSRIs) and/or psychological treatment with cognitive-behavioral therapy (CBT) (1). Although both treatments have proven to be effective (2, 3), they might have different neural working mechanisms. SSRIs are thought to work by dampening excessive limbic activity directly, while CBT targets fronto-limbic dysfunction indirectly via the engagement of dorsal prefrontal regions during cognitive therapy and directly via exposure and response prevention (4). Although there are several studies that have investigated the effects of these treatments on brain function, there is little direct evidence for this hypothesis.

A meta-analysis of resting cerebral blood flow and metabolism studies showed that pharmacological and psychological treatments reduce resting cortico–striato–thalamo–cortical circuit activity in OCD patients (5), circuits that are thought to play a key role in the pathophysiology of the disorder (6). A review of functional magnetic resonance imaging (fMRI) studies suggested that psychotherapy for OCD leads to decreased activity in ventral brain circuits during symptom provocation, as opposed to increased activity in the dorsal circuits during cognitive processing (7). Three studies have included both treatment types, but sample sizes were too small to compare these groups directly (n=4-9 per group) (8-10). A larger study that included both treatments used structural MRI, but neither performed a direct comparison between treatment groups (11). Therefore, the question whether treatment with CBT and SSRIs leads to similar or different functional changes in patients with OCD remains to be addressed.

We treated thirty-four OCD patients with sixteen weeks of SSRIs or CBT and compared pre- and post-treatment measures on task-based fMRI. We selected three different tasks, one to probe SSRI effects, one to probe CBT effects, and one to probe potential common effects related to symptom improvement. As SSRIs are known to reduce neural responses to emotional stimuli in the amygdala and related limbic structures in healthy volunteers and patients with major depressive disorder (12, 13), we selected an emotional face matching task that is known to activate the amygdala (14). As we presumed that CBT would lead to enhancement of top-down control of the prefrontal cortex, resulting in better inhibitory control over unwanted compulsive behaviors, we used a stop signal task (SST) engaging areas involved in response inhibition (15). To investigate potential common treatment effects, we included a symptom provocation task to target the underlying neural mechanisms of symptom improvement.

## Methods and Materials

### Participants

Forty-four patients with OCD were recruited from the outpatient clinic of the psychiatry department of the Amsterdam UMC and OCD expertise center of HSK group. Inclusion criteria were (1) age between 18-70 years; (2) a diagnosis of OCD according to DSM-IV; and (3) treatment indication with SSRIs and/or CBT. Exclusion criteria were (1) a diagnosis of bipolar disorder, current or past psychosis, primary alcohol or drug abuse; (2) a contraindication for MRI, such as metal implants, claustrophobia, and pregnancy; (3) major head trauma or neurological disease; (4) adequate treatment of OCD with a high dosed SSRI or CBT within 4 weeks before screening; and (5) current treatment with tricyclic antidepressant or antipsychotic medication. Drop-out was due to loss of contact or refusing the second scan (n=4), restriction of scanning due to COVID pandemic (n=2), possible pregnancy (n=2), side effects of medication (n=1) and premature ending of CBT protocol (n=1), resulting in data from 34 patients for analysis. Only three patients were taking a subtherapeutic SSRI dose at baseline (i.e. citalopram 20mg) (16). If patients preferred treatment with medication, the dose would be increased to therapeutic levels (n=1). In case of treatment with CBT, the medication would not be altered during the trial (n=2).

In addition, twenty healthy controls without a current or past psychiatric or neurological diagnosis and matched according to age, sex, and educational level, were recruited through flyers and online advertisements. All participants were assessed on the presence of psychiatric disorders using the M.I.N.I. International Neuropsychiatric Interview (17). The study was approved by the Ethical Committee of the Academic Medical Center in Amsterdam (METC 2016_127), registered at the Netherlands Trial Registry (NTR6575), and all participants provided written informed consent.

### Design and measurements

The study started as randomized controlled trial. As the majority of patients in the first months of the study did not consent with randomization, we continued the trial as a longitudinal non-randomised controlled trial in which patients were scanned prior to baseline and after 16 weeks of treatment (Figure 1). Choice of treatment was based on the preferences of patients and advise of treating psychiatrists. Treatment was performed according to the national Dutch guidelines (1), with either a high dosed SSRI or CBT sessions on a weekly basis. Pharmacological treatment consisted of citalopram 40-60mg (n=15) or sertraline 200mg (n=2) per day. CBT consisted of exposure with response prevention and cognitive therapy either in weekly group sessions at the AMC (n=13) or individual sessions at HSK group (n=4). Two patients that were treated with CBT used a stable subtherapeutic dose of citalopram during the therapy. Symptom severity was measured pre- and post-treatment using the Yale-Brown Obsessive-Compulsive Scale (Y-BOCS) (18), the Hamilton Anxiety Rating Scales (HAM-A) (19) and the Hamilton Depression Rating Scales (HAM-D) (20). Patients were defined as a treatment responder when they showed a reduction of ≥ 35% on the Y-BOCS (21). Twenty healthy controls (HC) were also scanned twice with a similar time interval to control for changes after naturalistic follow-up.

**Figure 1:**
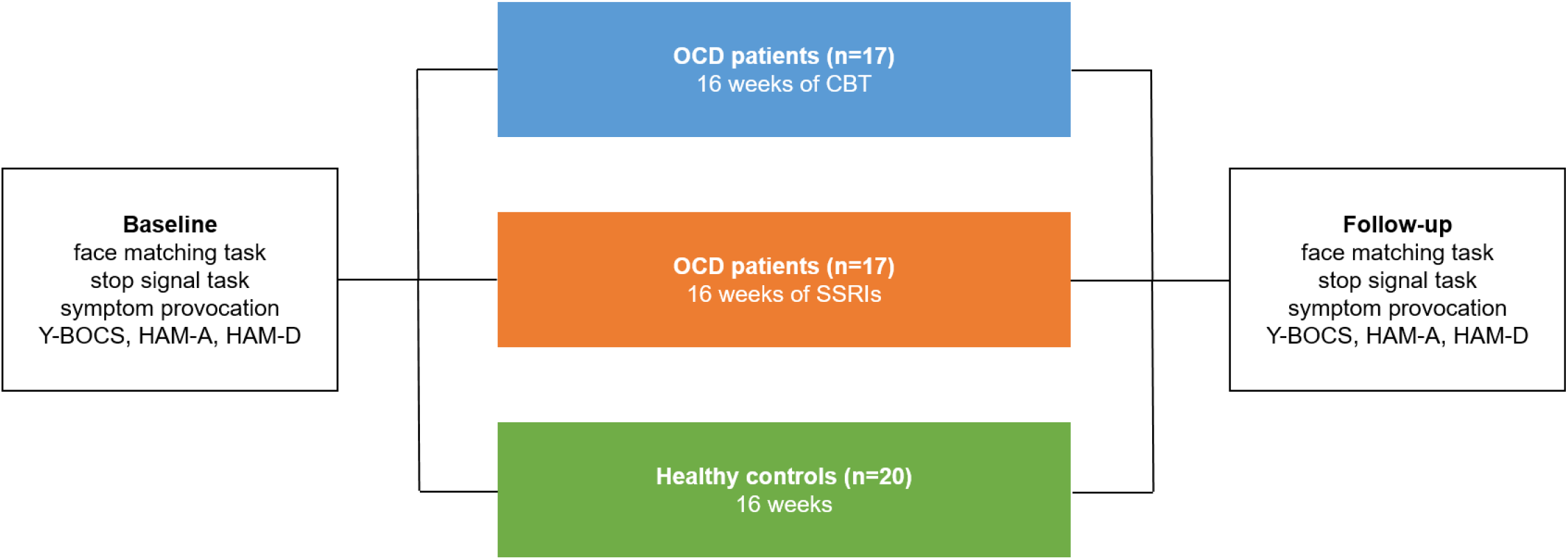
Study design. Functional magnetic resonance imaging was performed before and after treatment during emotional face matching, stop signal and symptom provocation tasks. Treatment consisted of either high dosed selective serotonin reuptake inhibitors (SSRIs) (n=17) or cognitive-behavioral therapy (CBT) sessions on a weekly basis (n=17). Twenty matched healthy controls were also scanned twice with a similar time interval. Scales: Yale-Brown Obsessive-Compulsive Scale (Y-BOCS), Hamilton Anxiety Rating Scale (HAM-A), Hamilton Depression Rating Scale (HAM-D).

### Neuroimaging tasks

Participants performed emotional face matching, stop signal and symptom provocation tasks at baseline and follow-up (see Supplemental Methods and Figures). The face matching paradigm consisted of a blocked design, including an emotional condition with angry and fearful faces and a control condition (14). The SST consisted of 180 go trials and 40 stop trials (15). Time between the onset of the go cue and the stop cue (stop signal delay; SD) was adjusted by a staircase procedure, resulting in an average inhibition success of approximately 50%. The symptom provocation task consisted of six blocks of three conditions with visual stimuli (i.e. OCD, fear and neutral). After every block participants were requested to rate how distressed they felt on a five-point scale. Three task versions were created with dimension specific stimuli (i.e. washing, checking or symmetry), so that patients were allocated to the stimuli sets related to their OCD subtype.

### Data acquisition

MR imaging was performed on a 3.0 T Philips MRI scanner, using a 32-channel SENSE head coil. The scanning protocol included a high-resolution T1-weighted MRI (sequence parameters: repetition time=6.9ms; echo time=3.1ms; flip angle=8°; 150 sagittal slices; voxel size=1.1mm isotropic, total duration=179s). For fMRI, multi-echo echoplanar imaging (EPI) was used to acquire T2*-weighted MRI volumes with blood oxygen level-dependent (BOLD) contrast (sequence parameters: repetition time=2375ms; number of echos=3, echo time=9, 26 and 44ms; flip angle=76°; field of view=224mm x 122mm x 224mm; voxel size=3.0 mm isotropic, 37 slices).

### Data analysis

Demographical, clinical and behavioral data were analyzed with SPSS (version 26.0). Group differences were tested using χ2 tests for categorical variables (gender, highest education, OCD subtype, presence of comorbidity and responder/non-responder). Continues variables were compared between groups with a one-way ANOVA (time between scans), Kruskal-Wallis test (age) or Welch’s t-test (duration of illness). The pre- and post-treatment scores on the Y-BOCS, HAM-A and HAM-D were compared using a repeated measures ANOVA with the factors group (SSRI, CBT) and time (baseline, follow-up) with post hoc t-testing in case of significant results with Bonferroni correction for multiple comparisons (alpha = 0.0125). The behavioral data of the SST were analyzed by calculating the accuracy on go and stop trials, reaction time (RT) on accurate go trials, stop signal reaction time (SSRT) and SSD (Supplemental Methods). The SSRT, SSD and RT on accurate go trials were compared using a repeated measures ANOVA with the factors group (SSRI, CBT, HC) and time (baseline, follow-up). Non-parametric tests were used to analyse the accuracy on go trials because the data follows a nonnormal distribution, including the Wilcoxon signed-rank test (main effect of time, averaged across groups) and Kruskal-Wallis test (main effect of group, averaged across time points). Mean RT and accuracy on the face matching task were calculated to check whether the participants performed the task according to the instructions. Validation of the symptom provocation task was assessed using the subjective distress ratings during the task. Because of the non-normal distribution, planned comparisons were carried out by dividing the analyses by group (Kruskal-Wallis test), and by conditions within groups (Friedman’s ANOVA), with Bonferroni correction for multiple comparisons (alpha = 0.0167). We additionally tested the main effect of time after summation of distress scores on all conditions and comparing these between time points (Wilcoxon signed-rank test).

### MRI analysis

MRI data were analyzed using SPM12 (www.fil.ion.ucl.ac.uk/spm/). First, realignment was performed and the three echoes were combined using in-house software to optimize BOLD contrast sensitivity and minimize signal dropout and distortion. Following co-registration of the mean EPI to the T1-weighted structural scan, functional images were normalized to Montreal Neurological Institute (MNI) space (voxel size of 2mm) and smoothed with a 3D Gaussian kernel of 8mm at full width at half maximum (FWHM). Participants with realignment parameters exceeding 3mm translation on the x-, y-, or z-axis were excluded from the analysis, resulting in the exclusion of the symptom provocation task of one patient. Due to scanner malfunction and lack of time, one patient did not complete the SST and one patient missed the face matching task. This resulted in the inclusion of thirty-three patients and twenty healthy controls per individual task.

After preprocessing, fMRI data were analyzed in the context of the general linear model. For the SST, the trials were modelled by convolving the onsets of the go stimuli with the canonical hemodynamic response function (HRF) for the following conditions: 1) correct go trials, 2) incorrect go trials, 3) successful stop trials and 4) failed stop trials. To probe brain regions involved in successful response inhibition, weighted contrasts were computed for each individual session for successful stop trials versus failed stop trials, which is seen as the most conservative approach (22). The face matching task was analyzed by modelling the two experimental conditions as box-car regressors convolved with the canonical HRF. Contrast images comparing the emotional face and visuo-motor control condition were obtained. The symptom provocation was analyzed by modelling three experimental conditions (OCD, fear, neutral). For the purpose of this study, we contrasted the OCD condition against the neutral condition. The realignment parameters were included into each model to account for movement artifacts, temporal autocorrelation was modeled using an autoregressive process, and a high-pass filter of 1/128 Hz was applied.

The contrast images of each task were entered into separate second-level group (SSRI, CBT, HC) × time (baseline, follow-up) interaction analyses to assess group differences in changes over time. We performed two planned Helmert contrasts to test common (1) and specific (2) brain changes, by 1) comparing the overall patient group and healthy controls over time and 2) comparing the two OCD treatment groups over time. Because CBT was more effective in treating OCD symptoms which resulted in a higher proportion of treatment responders, we added the pre and post Y-BOCS scores as covariates to the model for testing the second Helmert contrast (SSRI vs. CBT) to ensure that the reported results could not be explained by differences in symptom improvement. If the interaction analyses showed significant results we performed post-hoc t-testing to determine the direction of these results.

As we were interested in both common and divergent treatment effects we defined two regions of interest (ROI) per analysis: one general treatment ROI and one ROI targeting potential task specific differences. The general treatment ROI was based on a meta-analysis and systematic review looking into the treatment effects of OCD and included the bilateral caudate nucleus, thalamus and orbitofrontal cortex (OFC) (5, 7), based on their anatomical definition by the AAL atlas. In addition, task-specific ROIs were based on studies looking into baseline differences between OCD patients and controls on the three individual tasks. The face matching ROI consisted of the bilateral amygdala (23-26) and the symptom provocation ROI of the bilateral amygdala and insula (27, 28), both based on the AAL atlas. The SST ROI was defined as a sphere with a 10-mm radius around the coordinates of the left and right anterior cingulate and premotor cortex, areas that both showed abnormalities during inhibitory control in OCD in a recent meta-analysis (29).

Voxel-wise statistical tests were family-wise error (FWE) rate corrected (p<0.05) for multiple comparisons across the whole brain at the cluster level using a height threshold of p<0.001, or at peak level for the ROIs using a small volume correction (SVC, p<0.025, Bonferroni corrected for two ROIs per analysis).

## Results

### Demographic and clinical data

Demographic and clinical data are presented in Table 1. The three groups (SSRI, CBT and HC) showed no significant differences in age, gender, educational level and time interval between scanning sessions. Baseline Y-BOCS, HAM-A, HAM-D scores, the presence of comorbid diagnoses and OCD subtypes were not significantly different between SSRI and CBT groups (Supplemental Results). A group (CBT, SSRI) × time (baseline, follow-up) ANOVA revealed a main effect of time (F(1,32) = 71.72, p<0.001, η_p_^2^=0.69) with lower post-treatment Y-BOCS scores, a main effect of group (F(1,32) = 8.18, p=0.007, η_p_^2^=0.20) with lower averaged Y-BOCS scores in the CBT group and a group × time interaction (F(1,32) = 8.17, p=0.007, η_p_^2^=0.20) due to lower post-treatment Y-BOCS scores after CBT compared to SSRIs. The greater symptom reduction with CBT resulted in 76% responders after CBT and 24% responders after SSRIs. The group × time analysis of the other clinical scales only showed a main effect of time for the HAM-A (F(1,32) = 40.17, p<0.001, η_p_2=0.56) and HAM-D (F(1,32) = 27.80, p<0.001, η_p_^2^=0.47), with lower scores after treatment.

**TABLE 1.** Demographic and clinical data presented as the mean ± SD.

|  | SSRI (n=17) | CBT (n=17) | Controls (n=20) | p-value |
| --- | --- | --- | --- | --- |
| Age (years) | 30.06 ± 9.92 | 31.18 ± 8.56 | 29.90 ± 8.66 | p=0.704<sup>1</sup> |
| Gender (male/female) | 7/10 | 10/7 | 9/11 | p=0.553<sup>2</sup> |
| Highest education (n) |  |  |  | p=0.270<sup>2</sup> |
| – elementary school | 1 | 0 | 0 |  |
| – lower professional | 0 | 2 | 0 |  |
| – medium professional | 4 | 5 | 8 |  |
| – higher professional | 12 | 10 | 12 |  |
| Time between scans (weeks) | 18.45 ± 2.95 | 17.36 ± 6.18 | 17.34 ± 2.25 | p=0.653<sup>3</sup> |
| OCD subtype (n) |  |  |  | p=0.783<sup>2</sup> |
| – harm and checking | 6 | 9 |  |  |
| – contamination and washing | 3 | 3 |  |  |
| – symmetry and ordering | 4 | 3 |  |  |
| – forbidden thoughts | 3 | 1 |  |  |
| – miscellaneous | 1 | 1 |  |  |
| Duration of illness (years) | 12.88 ± 11.22 | 11.53 ± 9.60 | | p=0.712<sup>4</sup> |
| Comorbidity (n) |  |  |  | p=0.915<sup>2</sup> |
| – none | 5 | 5 |  |  |
| – single | 7 | 8 |  |  |
| – multiple ($\geq 2$) | 5 | 4 | | |
| Y-BOCS |  |  |  |  |
| – baseline | 25.82 ± 4.50 | 24.82 ± 3.13 | | p=0.458<sup>4</sup> |
| – follow-up | 19.65 ± 6.95 | 12.35 ± 5.86 | | p=0.002<sup>4*</sup> |
| Responder (yes/no) | 4/13 | 13/4 |  | p=0.002<sup>2*</sup> |
| HAM-A |  |  |  |  |
| – baseline | 18.71 ± 8.76 | 19.53 ± 8.55 | | p=0.783<sup>4</sup> |
| – follow-up | 11.47 ± 7.54 | 7.71 ± 7.54 | | p=0.155<sup>4</sup> |
| HAM-D |  |  |  |  |
| – baseline | 14.88 ± 5.76 | 12.47 ± 5.21 | | p=0.219<sup>4</sup> |
| – follow-up | 9.41 ± 5.87 | 5.94 ± 5.33 | | p=0.081<sup>4</sup> |

\* Significant difference between groups, $p \leq 0.05$ , tested with <sup>1</sup>Kruskal-Wallis test, <sup>2</sup>chi-square test, <sup>3</sup>one-way ANOVA or <sup>4</sup>Welch's t-test. Scales: Yale-Brown Obsessive-Compulsive Scale (Y-BOCS), Hamilton Anxiety Rating Scale (HAM-A), Hamilton Depression Rating Scale (HAM-D).

### Face matching task

The mean RT and accuracy indicated that the task was performed correctly (Table 2). The imaging analysis showed no baseline differences between patients and healthy controls for the face vs. neutral condition. First, we performed a group × time interaction analysis comparing both OCD groups with healthy controls, which showed no significant differences in changes over time. Second, the group × time interaction analysis with the two treatment groups (SSRI, CBT) revealed a significant cluster in the cerebellum (Figure 2A) and a cluster extending from the left posterior insula to Heschl’s gyrus (Figure 2B). Post-hoc paired-t testing showed that the interaction effect in the cerebellum was based on significantly increased activity after treatment in the CBT group (pFWE=0.021). The insula cluster also showed a relative increase in the CBT group and a relative decrease in the SSRI group, though these changes were not significant. We found no significant effects in the ROIs including the amygdala. All imaging results and statistics are presented in Table 3.

**TABLE 2.**
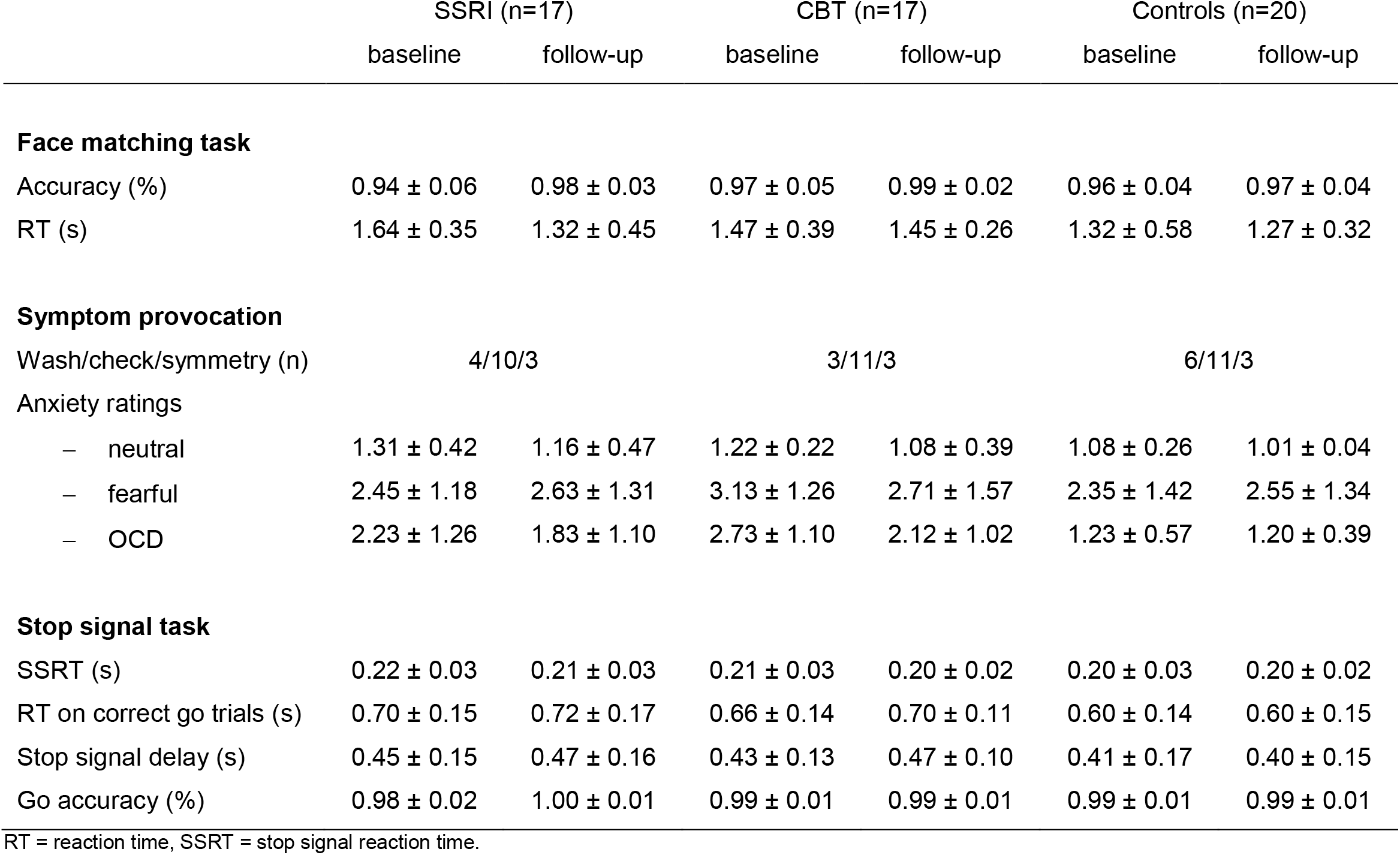
Task performance presented as the mean ± SD.

**TABLE 3.** Significant results from the interaction analyses to investigate common (group (OCD, HC) × time (baseline, follow-up)) and specific ((SSRI, CBT) × time (baseline, follow-up)) treatment effects.

| Contrast | MNI coordinates |  |  | Cluster size or Z-value | p-value | Region (AAL atlas) | BA | Direction of effect |
| --- | --- | --- | --- | --- | --- | --- | --- | --- |
|  | x | y | z |  |  |  |  |  |
| <u>Facematching task</u> |  |  |  |  |  |  |  |  |
| OCD vs. HC | - | - | - | - | - | - | - | - |
| SSRI vs. CBT | -2 | -52 | -6 | 520 | 0.001<sup>a</sup> | cerebellum | 18, 19 | ↑ CBT* |
|  | -42 | -18 | 8 | 249 | 0.026<sup>a</sup> | posterior insula –<br>heschl’s gyrus L | 48 | ↓ SSRI, ↑ CBT |
| <u>Stop signal task</u> |  |  |  |  |  |  |  |  |
| OCD vs. HC | - | - | - | - | - | - | - | - |
| SSRI vs. CBT | -24 | -14 | 30 | 362 | 0.006<sup>a</sup> | insula –<br>caudate nucleus L | 48 | ↑ CBT* |
|  | 18 | -86 | -6 | 235 | 0.034<sup>a</sup> | lingual gyrus R | 18, 19 | ↓ SSRI, ↑ CBT |
|  | -36 | 38 | -2 | 4.70 | 0.002<sup>b</sup> | orbitofrontal cortex L | 47 | ↓ SSRI, ↑ CBT |
|  | -26 | 10 | 60 | 4.06 | 0.013<sup>b</sup> | middle frontal gyrus L | 8 | ↓ SSRI, ↑ CBT |
| <u>Symptom provocation</u> |  |  |  |  |  |  |  |  |
| OCD vs. HC | 44 | 4 | -10 | 4.46 | 0.003<sup>b</sup> | insula R | 48 | ↓ in patients* |
| SSRI vs. CBT | 32 | -58 | 8 | 268 | 0.030<sup>a</sup> | calcerine sulcus –<br>hippocampus R | 19, 37 | ↑ SSRI, ↓ CBT |

<sup>a</sup> Whole-brain cluster-level correction. <sup>b</sup> SVC peak-level correction. \* Significant post-hoc paired t-test between pre- and post-treatment weighted contrasts. Groups: obsessive compulsive disorder (OCD), selective serotonin reuptake inhibitors (SSRI), cognitive behavioral therapy (CBT), healthy controls (HC). Direction of effect: ↑ increased activity, ↓ decreased activity.

**Figure 2:**
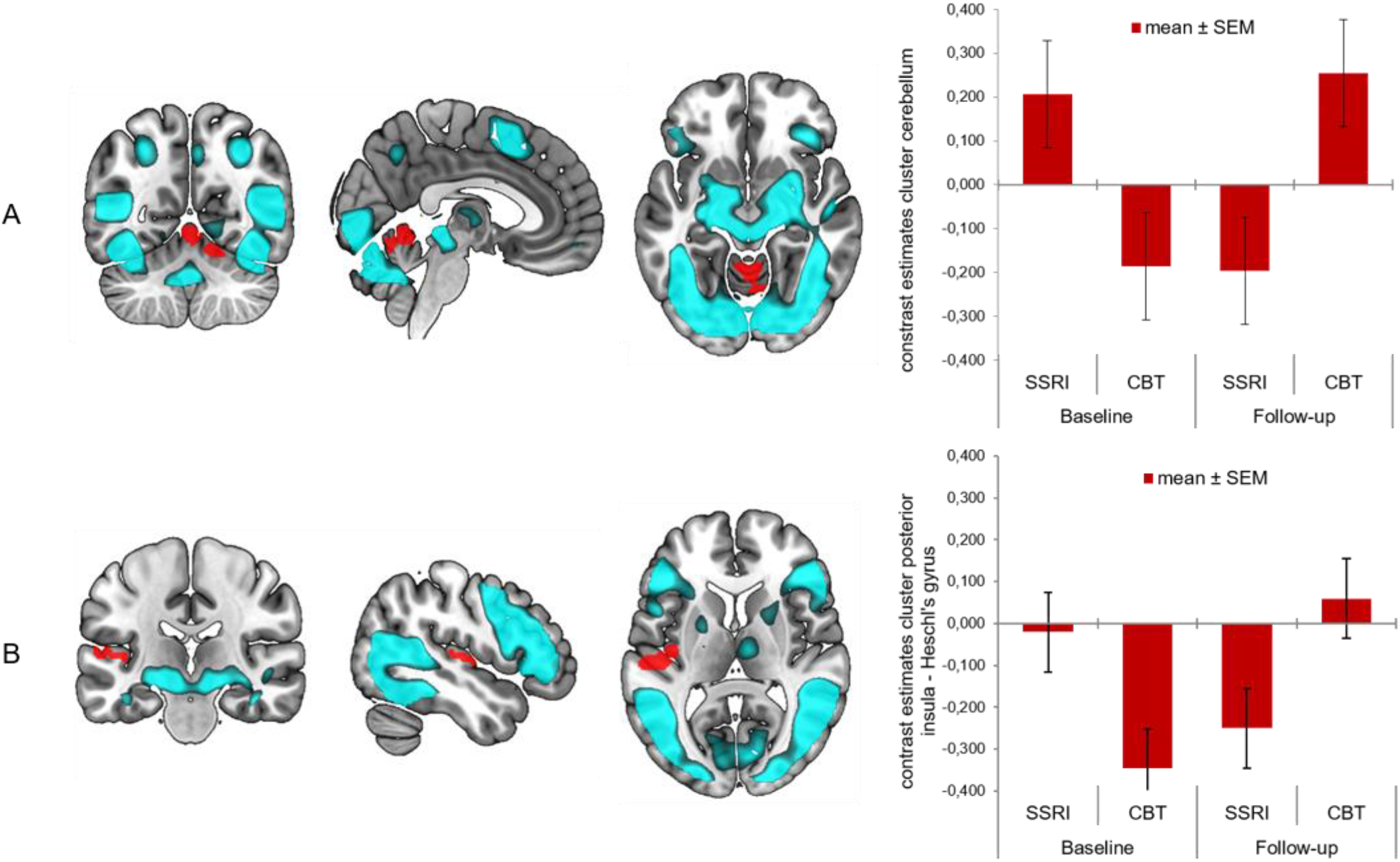
Results face matching task (face condition > neutral condition), with main effect of task (blue) and group by time analysis with patients treated with CBT compared to SSRIs (red); significant cluster in cerebellum (2A) and posterior insula – Heschl’s gyrus (2B).

### Stop signal task

The mean inhibition accuracy of 55% (SD = 4%) indicated that the staircase procedure was successful in balancing the number of successful and unsuccessful inhibition trials. Repeated measures ANOVA with the factors group (SSRI, CBT, HC) and time (baseline, follow-up) showed no significant differences for the SSRT, SSD and RT on accurate go trials. Likewise, the accuracy on go trials did not differ significantly between groups and over time.

We found no significant baseline imaging differences between patients and healthy controls for successful vs. failed stop trials. Similarly, the group × time interaction with all patients versus controls showed no significant differences in changes over time. The group × time interaction with the two treatment groups (SSRI, CBT) showed a significant cluster extending from the left insula to the caudate nucleus (Figure 3A) and a cluster in the right lingual gyrus (Figure 3B). Small volume correction for the predefined ROIs showed additional significant clusters in the left OFC (Figure 3C) and the left middle frontal gyrus (Figure 3D). Post-hoc paired-t testing showed that the interaction effect in the insula cluster was based on a significantly increased activity after CBT (pFWE<0.001). The other clusters also showed a relative increase in activity after CBT and a relative decrease in activity after SSRIs, but these changes were not significant.

**Figure 3:**
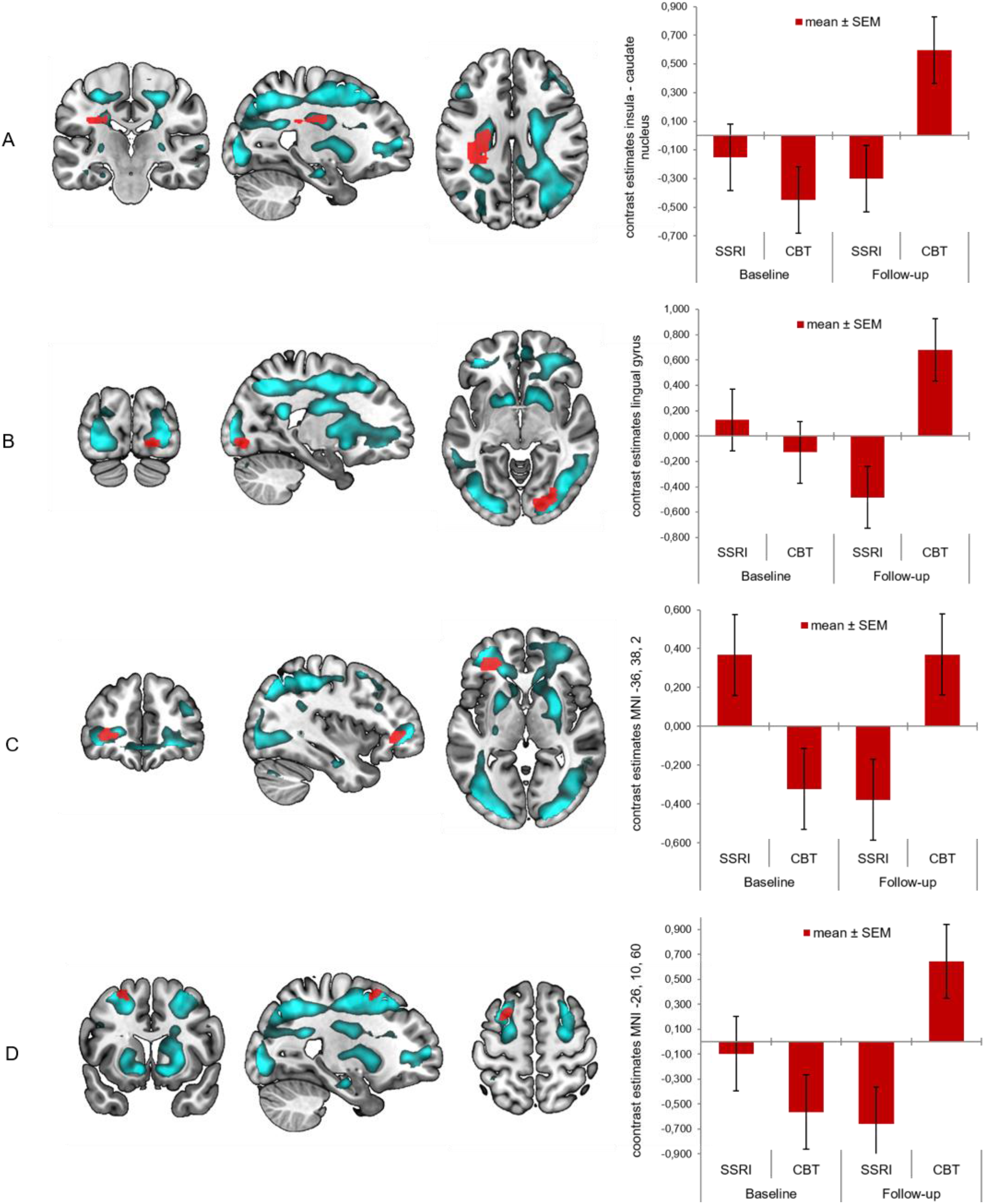
Results stop signal task (successful inhibition > failed inhibition), with main effect of task (blue) and group by time analysis with patients treated with CBT compared to SSRIs (red); significant cluster in insula to caudate nucleus (3A), lingual gyrus (3B), orbitofrontal cortex (3C) and middle frontal gyrus (3D).

### Symptom provocation task

Analyzing the subjective distress ratings showed that the symptom provocation was successful in inducing subjective distress in both treatment groups during the OCD and fear condition, while there was no main effect of time (Supplemental Results). We found no baseline imaging differences between patients and healthy controls for the OCD vs. neutral condition. The group × time interaction between both patient groups and controls showed a significant cluster in the right insula after small volume correction (Figure 4A). Post-hoc testing showed that this result was based on a decrease in activity after treatment in OCD patients (p(SVC)<0.001). The group × time interaction with the two treatment groups (SSRI, CBT) showed a significant cluster extending from the right calcarine sulcus to the hippocampus (Figure 4B). In contrast to the other results, this cluster showed a relative decrease in the CBT group and a relative increase in the SSRI group, but these changes were not significant.

**Figure 4:**
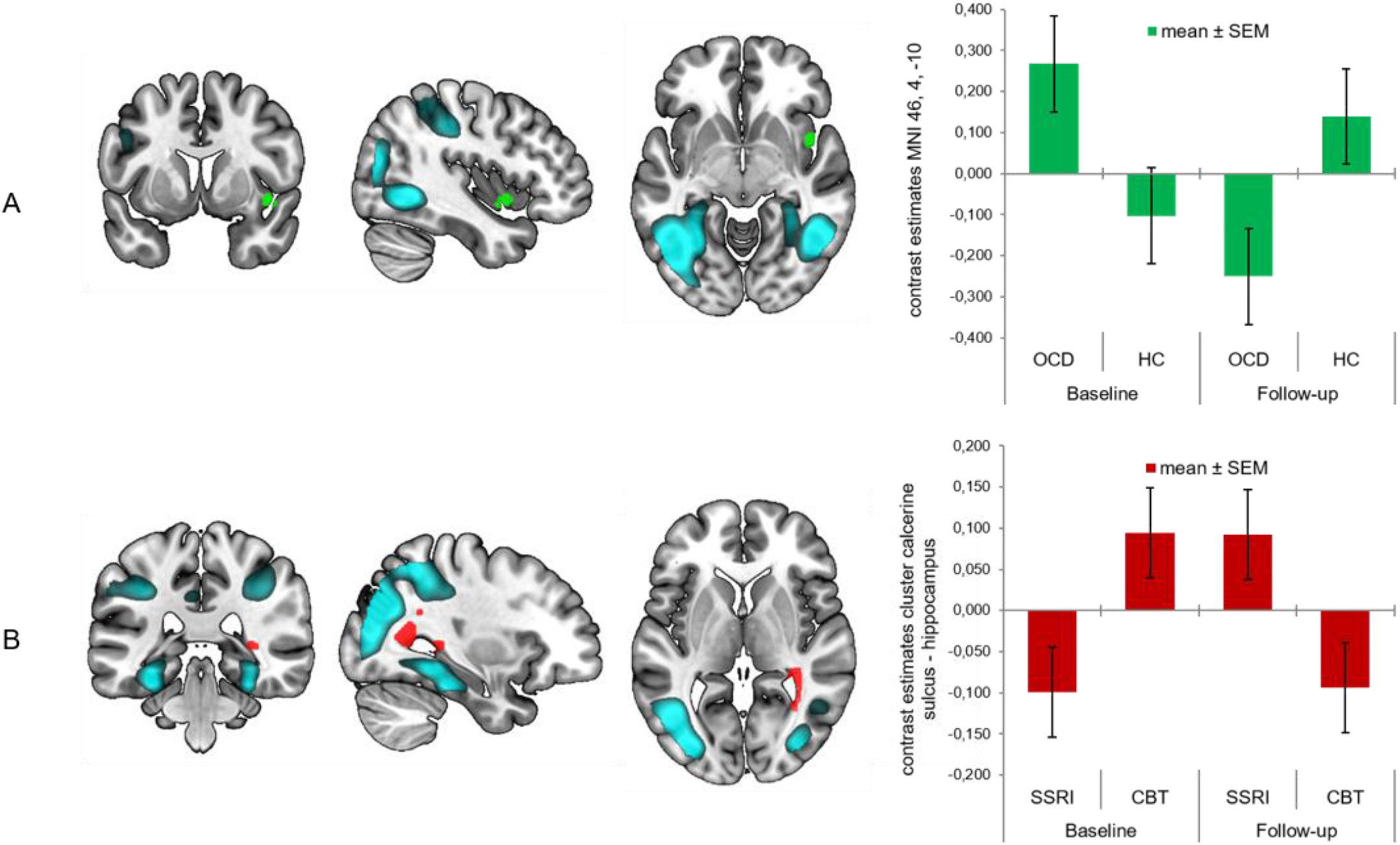
Results symptom provocation (OCD condition > neutral condition), with main effect of task (blue) and group by time analysis with OCD patients compared to healthy controls (green) and patients treated with CBT compared to SSRIs (red); significant cluster in insula (4A) and calcerine sulcus – hippocampus (4B).

### Sensitivity analysis

To ensure that these findings were not influenced by medication status, we reanalyzed the data after excluding the three patients that used medication at baseline. The analyses confirmed all of our main results, except the treatment induced changes in the posterior insula during face matching and the lingual gyrus during response inhibition.

## Discussion

This is the first study directly comparing the effects of pharmacological and psychological treatment on brain activity in OCD. Using task-based fMRI, we found that treatment with CBT and SSRIs leads to partly opposite functional changes in the brain, with a common reduction of insula activity during symptom provocation. CBT led to increased activity in the cerebellum and posterior insula during an emotional face matching task, while treatment with SSRIs resulted in relative decreases in these areas. Moreover, CBT led to a relative increase in activity in the lingual gyrus, OFC and middle frontal gyrus during response inhibition, compared to a relative decrease in these areas following SSRIs. Additionally, CBT significantly increased activity in a cluster extending from the caudate nucleus to the insula during response inhibition. Contrasting effects were observed in the right hippocampus during symptom provocation, with increased activity after SSRI treatment and decreased activity after CBT. The only common effect was observed during symptom provocation with reduced activity in the right insula after treatment in general. Together, these results suggest that psychological treatment works via increasing brain activity in various areas of the brain, while pharmacological treatment reduces activity in these same areas. Our results are not fully in line with the hypothesis that SSRIs work by dampening excessive limbic activity directly, while CBT targets fronto-limbic dysfunction indirectly via the engagement of dorsal prefrontal regions during cognitive therapy or directly via exposure and response prevention (4). Instead, we primarily observed opposite effects in cortical brain regions with increased activity after CBT and decreased activity after SSRIs, with little changes in limbic brain regions. Thus while CBT indeed appears to increase prefrontal engagement, we found little evidence for dampening of limbic activity by SSRIs. Our results therefore provide support for the generic hypothesis that CBT and SSRIs have distinct working mechanisms, but suggest those are primarily related to opposite cortical effects with little subcortical alterations. In fact, the differential treatment effects on the hippocampus during symptom provocation were even in the opposite direction, with increased activity after SSRIs and decreased activity after CBT. Together, our findings suggest that psychological treatment works via enhancement of compensatory cognitive mechanisms in various cortical brain areas, while pharmacological treatment dampens activity in these same areas through modulating neurotransmission.

In contrast to previous studies suggesting that brain changes following treatment were independent of treatment modality (8-10), our results suggest that most treatment effects are specific. We found significantly increased cerebellar activity during emotional processing after CBT. Evidence has emerged that the cerebellum is involved in emotional and cognitive functions well beyond motor control (30-34) and CBT has previously led to normalization of cerebellar function and structure in OCD patients (35-37). Our findings are located along the cerebellar vermis, branching out to a lateral region of the posterior cerebellum (lobule VI), which are often referred to as the ‘limbic cerebellum’. Lobule VI of the cerebellum is connected with the salience network, which plays a central role in detecting and filtering emotional stimuli and recruiting relevant functional networks (38). Evidence from lesion studies indicate that the vermis is involved in the formation of fear memory traces (39). Therefore, the cerebellar involvement might reflect the forming of new associations during CBT-related fear conditioning to facilitate appropriate responses to new situations (39). Although these interpretations remain preliminary, we suggest that involvement of the cerebellum is not only involved in the pathophysiology of OCD but also in CBT-induced recovery. During response inhibition, additional significant clusters were found in the lingual gyrus, OFC and the middle frontal gyrus. These results further support the hypothesis of CBT related neural engagement of frontal regions during response inhibition, possibly leading to better control over unwanted compulsive behaviors.

Although SSRIs showed the expected dampening of neural activity, we surprisingly found no treatment-induced changes in the amygdala during emotional processing, nor did we found baseline hyperactivity in OCD patients compared to healthy controls. It has been suggested that conflicting findings regarding involvement of the amygdala in OCD are the result of patient characteristics (i.e. medication status) and low statistical power (27). Our sensitivity analyses indicated that these results could not be explained by initial medication use. But although our sample size is large compared to previous small sample studies, our null finding could be the result of low statistical power.

In addition to the divergent effects, we found a common treatment effect consisting of decreased activity in the right insula during symptom provocation. The insula plays a role in emotional processing in OCD (27), and has been linked with OCD symptoms such as washing obsessions and disgust induction (40-43). Although previous studies concluded that normalization of the CSTC-circuit underlies treatment response (5), our results now suggest that the reduction of symptoms is also associated with decreased insular activity during emotional processing. This might be explained by the fact that previous studies mainly focused on resting cerebral activity, while we investigated the response to emotional stimuli.

This study has several limitations. First, the difference in response between treatments was larger than expected (3). This was mainly due to the high success rate of the intensive group CBT sessions (mean Y-BOCS difference = -14.08, SD = -6.78). Other possible explanations could be the sample size or the fact that the majority of patients refused randomization. The results should therefore be interpreted with caution. However, the analyses comparing both treatment groups were corrected for individual differences in symptom reduction by adding pre- and post-treatment Y-BOCS scores as a covariate to the analysis. The results therefore primarily reflect treatment effects rather than symptom improvement. Second, patients had various current or past comorbid disorders. Although this is a normal representation of the patient population, this might have affected our results. Third, longitudinal studies are vulnerable to selection biases as a result of loss to follow-up. We suspect that there was limited selective dropout while only two patients dropped out due to treatment-related factors (i.e. side effects of medication and premature ending of CBT protocol). Lastly, healthy controls were not screened for their family history of OCD, although previous research suggests that this could affect brain activity during response inhibition (15).

Despite these limitations, our results demonstrate mainly divergent but also common brain changes during emotional processing and inhibitory control, in response to psychotherapy and pharmacotherapy in OCD. These findings provide first insight in the common and specific neural mechanisms underlying treatment response, suggesting that CBT and SSRIs support recovery from OCD along partly distinct pathways.

## Data Availability

All data produced in the present study are available upon reasonable request to the authors

## Acknowledgements

This work was supported by the Netherlands Organization for Scientific Research (NWO/ZonMW Vidi 917.15.318). The article has been published as preprint on medRxiv.

