## Supplemental information for "Pharmacological and psychological treatment have common and specific effects on brain activity in obsessive-compulsive disorder"

**Supplemental Methods**

*Neuroimaging tasks*

Face matching

The paradigm consisted of a blocked design, including an emotional face condition and a control condition [1]. In the face condition, an angry or fearful face was presented on top as cue, and subjects had to indicate which of the bottom two faces matched the emotional expression of the top face by pressing the correct button. In the control condition, a horizontally or vertically oriented ellipse was presented on top as cue, and subjects had to indicate which of the bottom ellipses was identically oriented as the top ellipse. Two emotional face blocks were interleaved with three control blocks, and each block consisted of six 5 seconds trials, resulting in a total scan duration of 2.5 minutes (Supplementary Figure 1).

Stop signal

The SST consisted of 180 go trials and 40 stop trials with a total duration of ten minutes, inter-trial-intervals (ITI) were randomized between 400ms and 1500ms [2]. All stimuli were white schematics centered on a black background. Each trial started with a fixation plus sign (Supplementary Figure 2). In go trials, an arrow was shown in the center of the screen pointing either left or right. Participants were instructed to react as quickly as possible by pressing the button in congruence with the direction of the arrow. The same go cue was shown in stop trials, but was shortly followed by a stop cue in the form of an X, indicating that the participants should inhibit their response and not push any button. The time between the onset of the go cue and the stop cue (stop signal delay or SSD) was determined by a staircase procedure. The SSD started at 250ms and 50ms was either added or subtracted from the next stop trial based on the accuracy of their response. This way, task difficulty was negatively correlated with task performance, resulting in an average inhibition success of approximately 50%. All participants were told to react as fast as possible, but that accuracy on stop trials was equally important.

Symptom provocation

Participants underwent a symptom provocation task in which they were exposed to visual stimuli with OCD-related, general fear and neutral content (Supplementary Figure 3). The task was previously used in a study looking into the effect of psychological distress on stress-related circuitry in OCD (van Leeuwen et al., in prep). Three task versions were created with dimension specific stimuli, so that patients were allocated to the stimuli sets related to their OCD subtype (i.e. washing, checking or symmetry). Healthy controls were matched to these categories. The task contained 72 unique pictures in each session (baseline, follow-up), presented in 6 blocks of every condition (‘OCD’, ‘fear’ and ‘neutral’) in pseudo-random order. Each block started with 2 seconds fixation and consisted of 4 pictures, with a stimulus duration of 3.5 seconds and an inter-stimulus interval of 0.5 second, resulting in a total scan duration of 8 minutes. During the task participants were requested to envision themselves to the depicted scenes. After every block participants were requested to rate how anxious they felt on a five-point scale.

*Data analysis*

Stop signal

The behavioral data of the SST were analyzed by selecting 1) correct go trials, 2) incorrect go trials, 3) successful stop trials and 4) failed stop trials using matlab version R2017a (http://www.mathworks.com). We then calculated the accuracy on go trials, the accuracy on stop trials, the mean reaction time (RT) on correct go trials, the stop signal delay (SSD) and the stop signal reaction time (SSRT) for each individual session. The SSRT was calculated using the quantile method [3], by subtracting the SSD from the quantile reaction time (QRT). The QRT was obtained by sorting RTs on correct go trials ascendingly and calculating the RT corresponding to the quantile of the proportion of failed stop trials (which was approximately 50% for each participant). To ensure that participants were performing the task accurately, we restricted the proportion of inhibition failure and success (no more than 75% of stop trials) and correct go trials (no less than 60% of go trials), resulting in the exclusion of one patient that was treated with SSRIs. Afterwards, extreme outliers, i.e. data points that deviated more than three times the inter-quartile range, were removed from further testing in SPSS. The mean inhibition accuracy was calculated to check whether the staircase procedure worked in averaging inhibition success.

**Supplemental Results**

*Demographic and clinical data*

Patients had various current or past comorbid disorders at baseline, including generalized anxiety disorder (26%), major depressive disorder (21%), specific phobia (18%), hypochondriasis (18%), agoraphobia (9%), panic disorder (6%), dysthymic disorder (3%), social anxiety disorder (3%) and post-traumatic stress disorder (3%). OCD subtypes included harm and checking (44%), symmetry and ordering (21%), contamination and washing (18%), forbidden thoughts (12%) and miscellaneous (6%).

*Symptom provocation*

Validation of the symptom provocation was assessed using the subjective anxiety ratings during the task. The comparison between groups (SSRI, CBT, HC) showed a significant difference in averaged anxiety ratings (H(2) = 7.48, *p* = 0.024), with higher overall scores in the CBT group compared to the healthy controls. The Wilcoxon signed-rank test showed no main effect of time of the summed anxiety ratings across conditions (W=431.50, p=0.072). Planned comparisons within the three groups showed a main effect of condition for the SSRI group (*F*_r_ (2) = 19.96, *p*<0.001), the CBT group (*F*_r_ (2) = 17.77, *p*<0.001) and the healthy control group (*F*_r_ (2) = 29.93, *p* <0.001). Post-hoc testing showed that this was the result of higher anxiety ratings on both the OCD condition and fear condition compared to the neutral condition in the OCD groups, and higher anxiety scores on the fear condition compared to the OCD and neutral condition in the healthy control group. In summary, the symptom provocation was successful in inducing subjective anxiety in both treatment groups during the OCD and fear condition, while there was no main effect of time.

**Supplemental Figures**

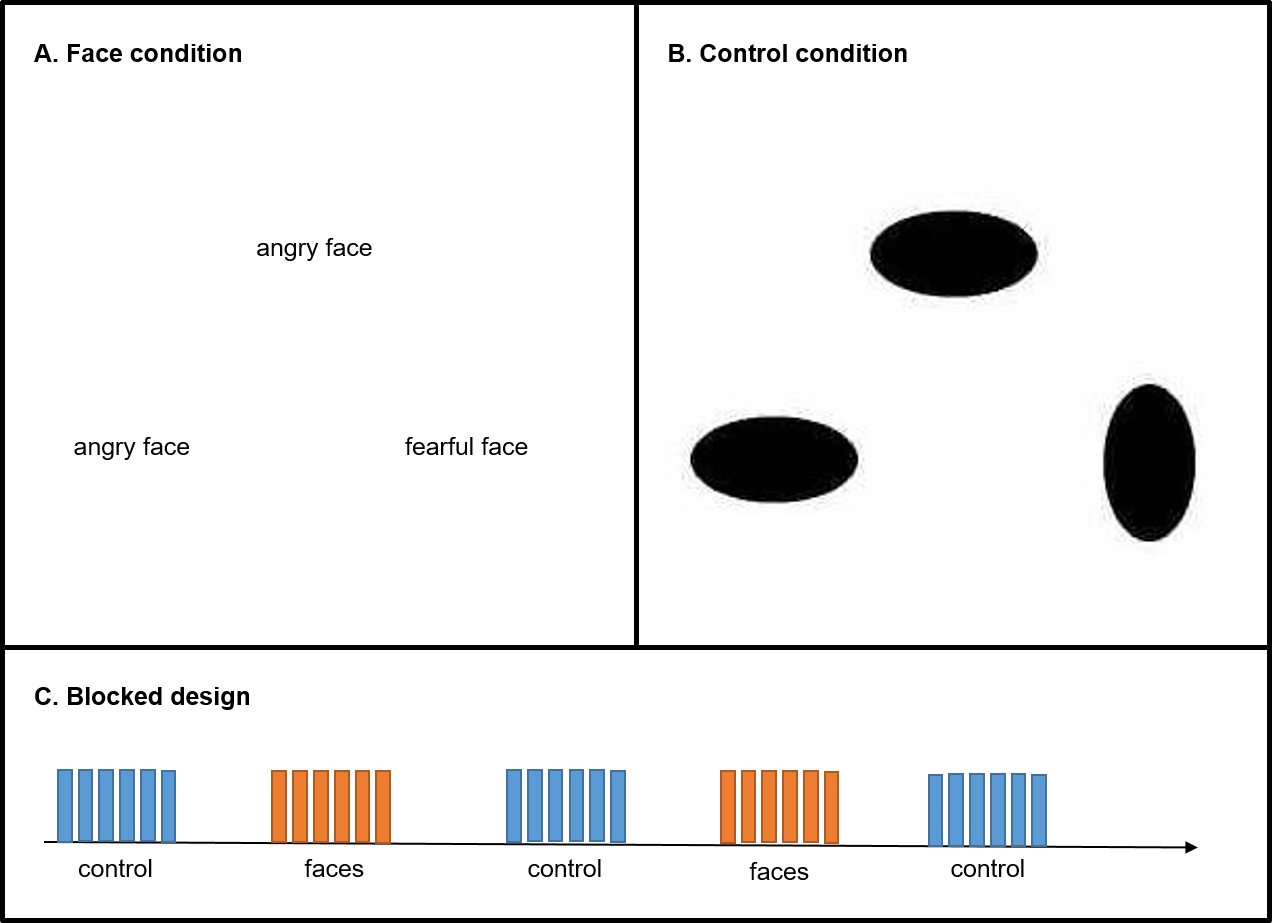

**Supplemental Figure 1: Face matching task A. Face condition.** An angry or fearful face was presented on top as cue, and subjects had to indicate which of the bottom two faces matched the emotional expression of the top face by pressing the correct button. **B. Control condition.** A horizontally or vertically oriented ellipse was presented on top as cue, and subjects had to indicate which of the bottom ellipses was identically oriented as the top ellipse by pressing the correct button. **C. Blocked design.** Two emotional face blocks were interleaved with three control blocks. Each block consisted of six 5 seconds trials, resulting in a total scan duration of 2.5 minutes.

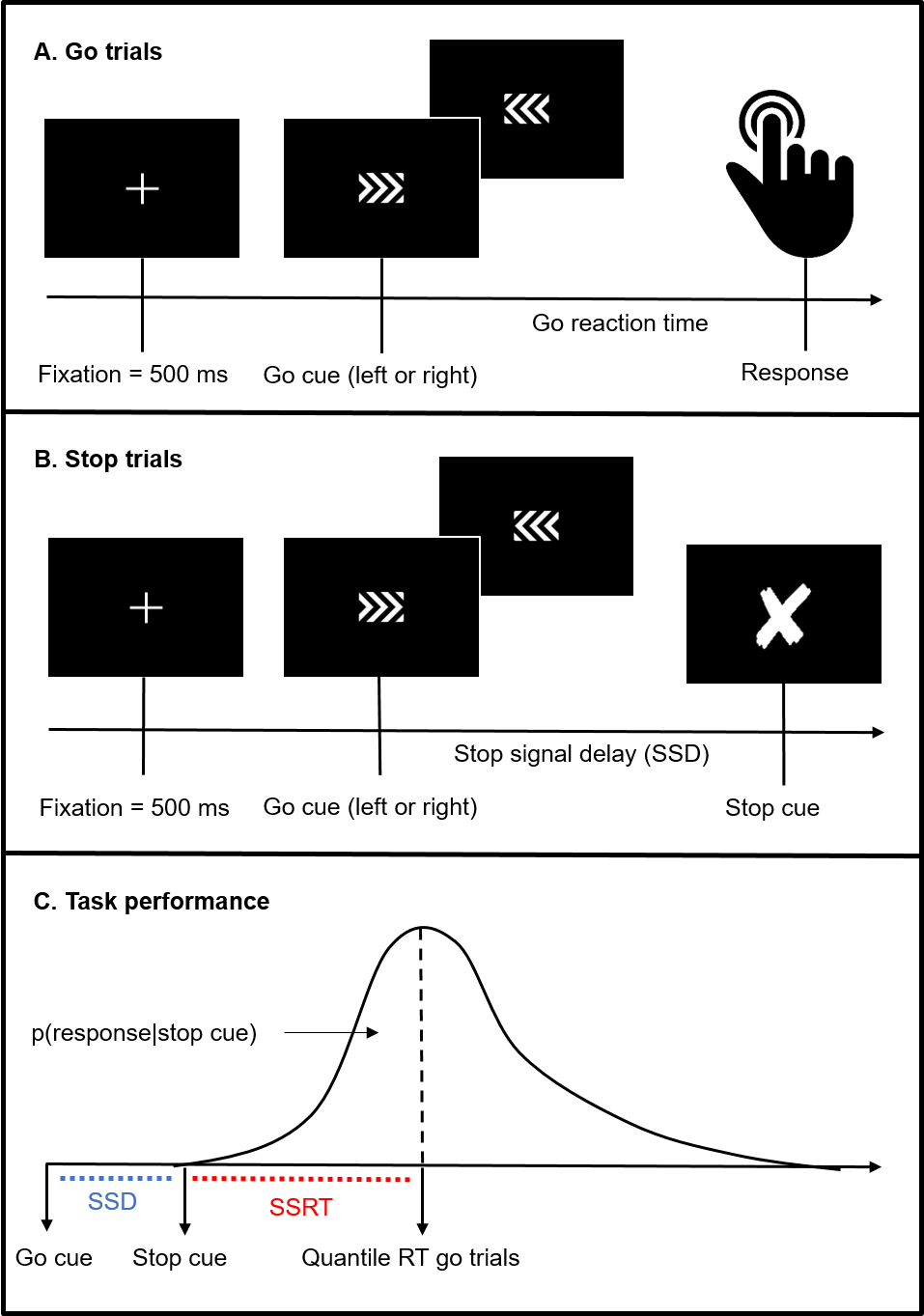

**Supplemental Figure 2: Stop Signal Task A. Go-trials.** After a fixation cue, a go-cue is presented to which patients have to react by pressing a button. They either succeed (successful go trial) or fail (failed go trial) and reaction times are measured (go reaction time). **B. Stop-trials.** After the go-cue a stop-cue is presented indicating that a response has to be withheld. The stop-signal delay (SSD) is varied using a staircase procedure to make sure that participants have an inhibition success of approximately 50%. Participants either succeed (successful inhibition) or fail (failed inhibition). **C. Task performance.** Stop-signal reaction time (SSRT) is calculated by subtracting the mean SSD from the quantile reaction time (QRT). The QRT was obtained by sorting RTs on correct go trials ascendingly and calculating the RT corresponding to the quantile of the proportion of failed stop trials (which was approximately 50% for each participant).

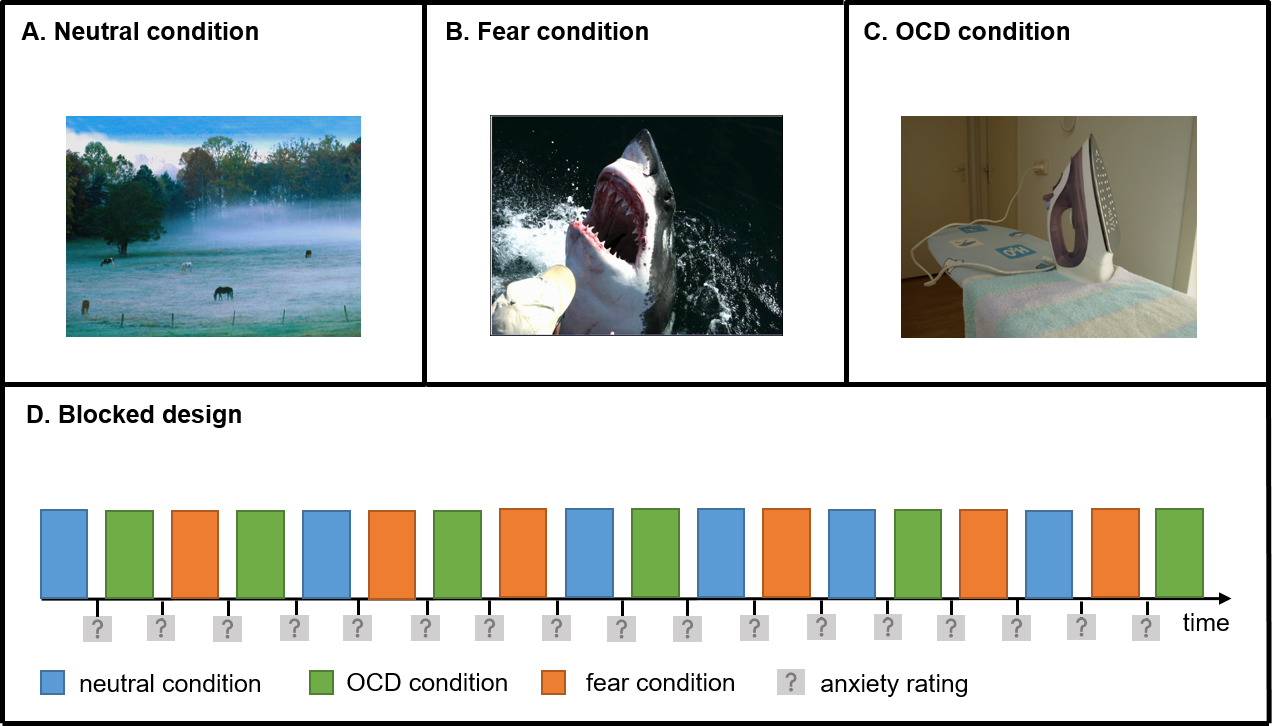

**Supplemental Figure 3: Symptom provocation task. A. Neutral condition; B. Fear condition; C. OCD condition.** Participants underwent a symptom provocation task in which they were exposed to 72 unique OCD-related, fear and neutral pictures. Three task versions were created with OCD dimension specific stimuli (i.e. washing, checking or symmetry). **D. Blocked design.** The task consisted of 6 blocks of every condition (neutral, fear, OCD). Each block started with 2 seconds fixation and consisted of 4 pictures with a stimulus duration of 3.5 seconds and an inter-stimulus interval of 0.5 second, resulting in a total scan duration of 8 minutes. After every block participants were requested to rate how anxious they felt on a five-point scale.
